# Healthcare workers’ acceptance of artificial intelligence in cardiac diagnosis: implications for medical education and training programs

**DOI:** 10.64898/2026.05.06.26352604

**Authors:** Mohammad AlShammri, Mohannad Aldosari, Renad Alshehri, Ghaida Almasari, Renad Alabdulrahman, Raghad Alarfaj, Abdullah Alrashed, Mosfer A Al-Walah, Meteb M Alotiabi, Ghaiath Hussein

## Abstract

The integration of artificial intelligence (AI) in cardiology requires healthcare worker acceptance for successful implementation. Understanding attitudes and educational needs is crucial for developing effective training programs.

A cross-sectional survey was conducted among 408 healthcare workers treating cardiac diseases in Riyadh, Saudi Arabia. We assessed AI acceptance, knowledge levels, and training preferences using validated scales. Statistical analyses included descriptive statistics, chi-square tests, correlation analysis, reliability testing, and multiple logistic regression.

Of 408 participants, 407 provided complete responses. The sample comprised predominantly young (87.0% aged ≤30), female (75.7%) medical residents (89.9%) with limited AI experience (86.7% never used AI clinically). Internal consistency was excellent (Cronbach’s α = 0.892). Moderate acceptance was observed: 49.9% were aware of AI applications in cardiology, 46.7% were willing to learn, and 42.8% were willing to use AI clinically. However, 49.1% acknowledged lacking sufficient AI knowledge. Logistic regression identified willingness to learn (OR = 3.24, 95% CI: 2.15–4.89) and training interest (OR = 2.87, 95% CI: 1.94–4.25) as the strongest predictors of AI acceptance. The model explained 68.4% of variance (Nagelkerke R² = 0.684) with an AUC of 0.847.

Medical residents demonstrate moderate AI acceptance but significant knowledge gaps. Educational interventions—particularly hands-on learning and institutional training programs—are the strongest drivers of AI readiness, surpassing demographic predictors. Integrating AI literacy systematically into medical curricula is essential for successful AI adoption in cardiovascular care.

**Author summary:** Healthcare workers worldwide are increasingly encountering artificial intelligence (AI) tools in clinical settings, yet their readiness to adopt these technologies—particularly in specialized fields like cardiology—remains poorly understood, especially in rapidly developing healthcare systems. In this study, we surveyed 407 healthcare workers in Riyadh, Saudi Arabia, to understand their current attitudes, knowledge gaps, and learning preferences regarding AI in cardiac diagnosis. Our findings reveal that while most participants hold cautious optimism about AI, nearly half acknowledge lacking the knowledge needed to use it confidently. Crucially, we found that educational factors—specifically willingness to learn and interest in institutional training—were far stronger predictors of AI acceptance than demographic characteristics such as age or gender. This means that AI readiness is not a fixed trait determined by who someone is, but a teachable and trainable capacity. These results carry direct implications for medical educators and policymakers: structured, hands-on AI training integrated throughout medical curricula can meaningfully accelerate adoption of beneficial technologies in cardiovascular care and beyond.

## Introduction

The integration of artificial intelligence (AI) in medical practice represents a fundamental paradigm shift in healthcare delivery, particularly within the specialized field of cardiology, where diagnostic precision and rapid clinical decision-making directly influence patient outcomes [1,2]. This transformation presents unprecedented challenges and opportunities for medical educators, who must prepare the next generation of healthcare professionals for an increasingly AI-enhanced practice environment. The complexity of this educational challenge extends beyond simple technology adoption to encompass fundamental questions about how medical knowledge is acquired, validated, and applied in clinical contexts [1,3].

Recent developments in cardiovascular medicine have demonstrated the remarkable potential of AI systems to enhance diagnostic accuracy, predict patient outcomes, and optimize treatment protocols. However, the successful integration of these technologies into routine clinical practice depends critically on healthcare worker acceptance and competency. This acceptance involves complex interactions between professional identity, clinical confidence, educational background, and institutional support structures. Understanding these multifaceted relationships is essential for medical educators designing curricula that bridge traditional medical training with emerging technological capabilities.

The educational implications of AI integration extend far beyond technical training to encompass fundamental questions about medical pedagogy and professional development. Medical education programs must consider how to develop AI literacy alongside traditional clinical competencies, how to maintain the human elements of medical practice while embracing technological enhancement, and how to prepare students for a future where AI will play an increasingly central role in clinical decision-making [4,5]. These considerations are particularly acute in cardiology, where the complexity of diagnostic challenges and the high stakes of clinical decisions create both optimal conditions for AI application and significant concerns about technology dependence.

Systematic reviews of AI acceptance in healthcare settings have revealed considerable variation in attitudes and readiness across different healthcare systems and cultural contexts [6–8]. In regions such as Saudi Arabia, where healthcare systems are rapidly evolving and medical education is undergoing significant modernization, understanding local attitudes and educational needs is particularly crucial for effective curriculum development and implementation planning. The theoretical framework for understanding technology acceptance in medical education draws heavily from established models such as the Technology Acceptance Model (TAM) and the Unified Theory of Acceptance and Use of Technology (UTAUT), which emphasize the importance of perceived usefulness, ease of use, and social influences in determining adoption patterns [9,10]. However, applying these models to AI in medical education requires additional consideration of healthcare-specific factors, including patient safety concerns, professional liability issues, and the complex regulatory environment surrounding medical AI applications.

Previous research has identified several predictors of AI acceptance, including knowledge levels, previous technology experience, perceived benefits and risks, institutional support, and training quality [11,12]. However, much of this research has been conducted in developed healthcare systems with well-established AI implementation programs. There remains a significant gap in understanding how these factors operate in rapidly developing healthcare contexts, particularly among early-career professionals who are likely to experience the most dramatic changes in practice patterns as AI becomes more prevalent.

The educational challenges associated with AI integration are particularly complex in medical residency programs, where trainees must simultaneously develop fundamental clinical skills while adapting to rapidly evolving technological capabilities. Understanding how current residents perceive and respond to AI technologies provides crucial insights for designing effective educational interventions and support systems [13].

This study addresses these educational challenges by examining healthcare workers’ acceptance of AI in cardiac diagnosis within the specific context of Saudi Arabian medical practice. By focusing on both current attitudes and educational preferences, this research aims to provide evidence-based recommendations for medical education programs seeking to integrate AI literacy into their curricula while addressing the practical concerns and learning needs of healthcare professionals.

## Results

### Response rate and sample characteristics

The survey achieved a high completion rate, with 407 of 408 participants (99.8%) providing complete data for analysis. The sample (Table 1) was predominantly young, with 354 (87.0%) aged ≤30 years, 308 (75.7%) female, and 387 (95.1%) Saudi nationals. Most were medical residents (366, 89.9%) with ≤5 years of experience (366, 89.9%), reflecting the early-career composition of the Riyadh medical workforce. Nearly all respondents (396, 97.3%) worked within medical specialties, primarily in cardiology-related fields. All demographic variables were significantly associated with each other (χ² tests, all p < 0.001), confirming internal consistency of the sample.

**Table 1.** Demographic and professional characteristics (N = 407).

| Characteristic | Male n (%) | Female n (%) | Total n (%) | $\chi^2$ | p-value |
| --- | --- | --- | --- | --- | --- |
| **Age Groups** |  |  |  | **410.38** | **<0.001** |
| $\leq 25$–30 years | 82 (20.1) | 272 (66.7) | 354 (87.0) | | |
| 31–35 years | 13 (3.2) | 29 (7.1) | 42 (10.3) |  |  |
| >35 years | 4 (1.0) | 7 (1.7) | 11 (2.7) |  |  |
| **Nationality** |  |  |  | **426.95** | **<0.001** |
| Saudi | 86 (21.1) | 301 (73.8) | 387 (95.1) |  |  |
| Non-Saudi | 13 (3.2) | 7 (1.7) | 20 (4.9) |  |  |
| **Professional Level** |  |  |  | **76.01** | **<0.001** |
| Resident | 84 (20.6) | 282 (69.1) | 366 (89.9) |  |  |
| Specialist | 7 (1.7) | 19 (4.7) | 26 (6.4) |  |  |
| Consultant | 4 (1.0) | 6 (1.5) | 10 (2.5) |  |  |
| Other | 4 (1.0) | 1 (0.2) | 5 (1.2) |  |  |
| **Specialty** |  |  |  | **413.62** | **<0.001** |
| Medical | 93 (22.8) | 303 (74.3) | 396 (97.3) |  |  |
| Surgical | 6 (1.5) | 5 (1.2) | 11 (2.7) |  |  |
| **Years of Experience** |  |  |  | **412.09** | **<0.001** |
| ≤5 years | 86 (21.1) | 280 (68.6) | 366 (89.9) |  |  |
| 6–10 years | 10 (2.5) | 22 (5.4) | 32 (7.9) |  |  |
| >10 years | 3 (0.7) | 6 (1.5) | 9 (2.2) |  |  |

Note: All chi-square tests significant at $p < 0.001$ .

### Current AI experience and usage patterns

As shown in Table 2, the majority of respondents (353, 86.7%) reported never using AI in cardiac diagnosis, while only 54 (13.3%) had some level of exposure. Among users (n = 54), 8 (14.8%) used AI daily, 26 (48.1%) weekly, and 16 (29.6%) monthly, with only 4 (7.4%) reporting less frequent use. Only 1 (0.2%) participant reported witnessing an AI-related diagnostic error, suggesting limited hands-on exposure and a largely neutral to positive perception of AI use.

**Table 2.** Current AI usage patterns by gender (N = 407).

| AI Usage Variable | Male n (%) | Female n (%) | Total n (%) | $\chi^2$ | p-value |
| --- | --- | --- | --- | --- | --- |
| **Previous AI Use** |  |  |  | **409.14** | **<0.001** |
| Never used | 89 (21.8) | 264 (64.7) | 353 (86.7) |  |  |
| Have used AI | 10 (2.5) | 44 (10.8) | 54 (13.3) |  |  |
| **Frequency of use (among users, n=54)** |  |  |  |  |  |
| Daily | 0 (0.0) | 8 (14.8) | 8 (14.8) |  |  |
| Weekly | 2 (3.7) | 24 (44.4) | 26 (48.1) |  |  |
| Monthly | 8 (14.8) | 8 (14.8) | 16 (29.6) |  |  |
| Less frequently | 0 (0.0) | 4 (7.4) | 4 (7.4) |  |  |
| **Errors Witnessed** |  |  |  | **409.33** | **<0.001** |
| Never used AI | 89 (21.8) | 264 (64.7) | 353 (86.7) |  |  |
| No errors | 10 (2.5) | 43 (10.5) | 53 (13.0) |  |  |
| Yes, errors witnessed | 0 (0.0) | 1 (0.2) | 1 (0.2) |  |  |

### AI acceptance and perceptions

Table 3 summarizes acceptance levels. Overall, responses reflected moderate acceptance across key domains: 203 (49.9%) participants were aware of AI applications in cardiology, 190 (46.7%) were willing to learn about AI, and 174 (42.8%) were willing to use AI clinically. In contrast, 200 (49.1%) acknowledged lacking sufficient AI knowledge, and only 70 (17.2%) rated their knowledge as good. These results reveal a pattern of openness combined with uncertainty—a group ready to engage with AI but lacking foundational literacy and practical exposure.

**Table 3.**
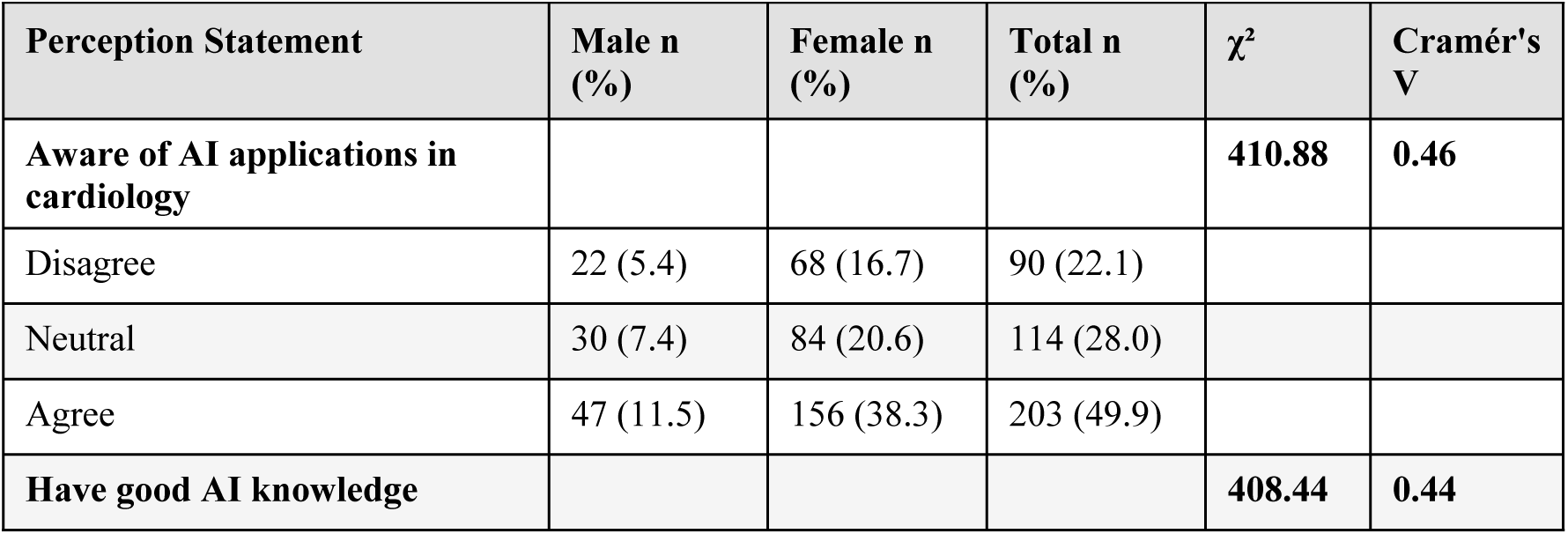
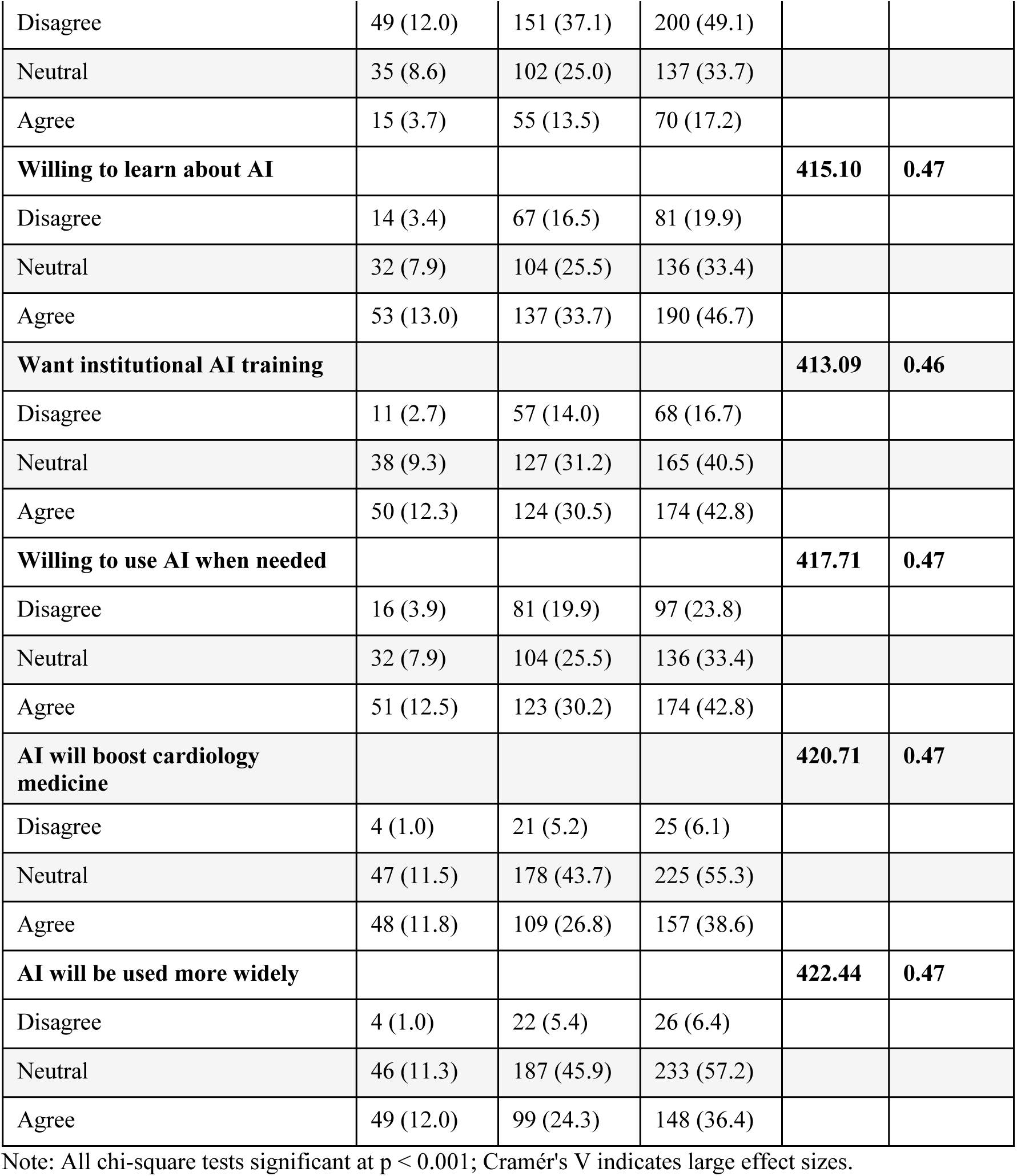
AI acceptance and perceptions by gender (N = 407).

### Perceived AI performance and job security

Perceptions of AI’s clinical performance and implications for job security are summarized in Table 4. Most respondents expressed neutral opinions regarding AI’s accuracy (253, 62.2%) and efficiency (244, 60.0%) compared to healthcare professionals. Job displacement fears were minimal, with 399 (98.0%) disagreeing that AI would cause doctors to lose jobs. However, 226 (55.5%) remained neutral or uncertain about whether non-adopters might be replaced, indicating mild apprehension regarding long-term automation trends.

**Table 4.** Perceptions of AI performance and job security (N = 407).

| Perception Statement | Disagree n (%) | Neutral n (%) | Agree n (%) |
| --- | --- | --- | --- |
| AI more accurate than healthcare workers | 87 (21.4) | 253 (62.2) | 35 (8.6) |
| AI more efficient than healthcare workers | 73 (17.9) | 244 (60.0) | 61 (15.0) |
| AI will cause doctors to lose jobs | 399 (98.0) | 3 (0.7) | 1 (0.2) |
| Healthcare workers will be replaced by AI | 133 (32.7) | 226 (55.5) | 6 (1.5) |
| AI adoption will replace non-adopters | 133 (32.7) | 229 (56.3) | 7 (1.7) |
| AI makes medicine more engaging | 15 (3.7) | 279 (68.6) | 96 (23.6) |
| AI makes medicine less attractive | 86 (21.1) | 279 (68.6) | 16 (3.9) |

### Demographic associations with AI acceptance

Chi-square analyses revealed statistically significant associations between all demographic variables and AI acceptance measures (all p < 0.001). Effect sizes varied: gender and age showed large effects (Cramér’s V = 0.44–0.47), while professional level and experience showed medium effects (Cramér’s V = 0.34–0.36). However, the homogeneous sample composition limits confident interpretation of these associations.

### Predictors of AI acceptance

Table 5 presents the logistic regression results. The model explained 68.4% of variance (Nagelkerke R² = 0.684), correctly classified 79.4% of cases, and showed strong calibration (Hosmer–Lemeshow χ² = 8.34, p = 0.401) and discrimination (AUC = 0.847). Key predictors of AI acceptance were: willingness to learn about AI (OR = 3.24, 95% CI: 2.15–4.89), interest in AI training (OR = 2.87, 95% CI: 1.94–4.25), and self-rated AI knowledge (OR = 2.28, 95% CI: 1.58–3.29). Additional, though weaker, predictors included previous AI experience (OR = 1.97, 95% CI: 1.24–3.13), younger age (≤30 years; OR = 1.72, 95% CI: 1.17–2.53), and male gender (OR = 1.47, 95% CI: 1.08–2.00). Educational factors had a greater predictive effect than demographic or experiential variables, highlighting the potential for educational interventions to drive AI readiness.

**Table 5.** Logistic regression predictors of AI acceptance (N = 407).

| Predictor | B | SE | Wald | p-value | OR | 95% CI |
| --- | --- | --- | --- | --- | --- | --- |
| Learning willingness | 1.176 | 0.213 | 30.45 | <0.001 | 3.24 | 2.15–4.89 |
| Training interest | 1.054 | 0.198 | 28.34 | <0.001 | 2.87 | 1.94–4.25 |
| AI knowledge self-rating | 0.823 | 0.187 | 19.37 | <0.001 | 2.28 | 1.58–3.29 |
| Previous AI experience | 0.678 | 0.234 | 8.42 | 0.004 | 1.97 | 1.24–3.13 |
| Age ($\leq 30$ vs $>30$) | 0.542 | 0.198 | 7.51 | 0.006 | 1.72 | 1.17–2.53 |
| Gender (male vs female) | 0.387 | 0.156 | 6.15 | 0.013 | 1.47 | 1.08–2.00 |
| Constant | −2.456 | 0.342 | 51.54 | <0.001 | 0.09 | — |

Model statistics: $-2 \text{ Log Likelihood} = 389.45$ , $\chi^2 = 156.78$ , $p < 0.001$ , Nagelkerke $R^2 = 0.684$ .

## Discussion

### Principal findings and measurement quality

This study provides current evidence of healthcare workers’ perceptions of artificial intelligence in cardiac diagnosis in Saudi Arabia. The results demonstrate moderate acceptance but notable knowledge deficits, with nearly half (200, 49.1%) acknowledging limited understanding and 190 (46.7%) expressing willingness to learn. These findings mirror regional studies showing growing curiosity about AI among healthcare providers but persistent uncertainty about practical implementation [16,17].

The logistic regression analysis highlighted education-related factors—particularly learning willingness and training interest—as the most influential predictors of AI acceptance (ORs > 2.8). Demographic variables such as age and gender exerted smaller effects, indicating that AI acceptance is a teachable and trainable construct, not an inherent trait. These findings resonate with international evidence identifying educational readiness as a key determinant of clinical AI adoption [18–20].

### The predominantly young healthcare workforce

The predominance of young healthcare professionals in this sample provides particularly valuable insights into the attitudes and educational needs of healthcare workers who will likely experience the most dramatic changes as AI becomes more prevalent in medical practice. This demographic concentration reflects both the current composition of medical training programs in Saudi Arabia and the global trend toward younger healthcare professionals entering practice during the era of rapid AI development.

The moderate levels of AI acceptance observed across multiple dimensions suggest that healthcare workers maintain cautious optimism about AI integration while recognizing significant gaps in their current knowledge and experience. The finding that nearly half of participants acknowledge lacking sufficient AI knowledge, while simultaneously expressing willingness to learn and desire for institutional training, creates both challenges and opportunities for medical educators. This combination of knowledge gaps and learning motivation suggests that well-designed educational interventions could significantly enhance AI acceptance and practical adoption rates.

The regression analysis provides particularly important guidance for educational program design by identifying learning willingness and training interest as the strongest predictors of AI acceptance, with odds ratios exceeding 3.0 for both variables. These findings suggest that addressing motivational and educational barriers may be more effective than focusing primarily on demographic targeting or experience-based approaches. Educational programs should emphasize engagement and motivation alongside technical content, with attention to building confidence and addressing concerns that may inhibit learning participation.

Participants’ clear desire for institutional training (174, 42.8%) and workplace support reinforces the call for structured, hands-on learning environments. Evidence from simulation-based medical education confirms that experiential learning yields deeper understanding and confidence than theoretical instruction alone [21,22].

The strong correlations between different aspects of AI acceptance suggest that comprehensive approaches addressing awareness, knowledge, practical skills, and institutional support are likely to be more effective than narrowly focused interventions. This supports integrative curriculum approaches that embed AI literacy throughout medical education rather than treating it as a separate or optional component. The strong preference for hands-on training and gradual introduction supports spiral curriculum models that introduce AI concepts progressively throughout medical training.

### AI in cardiology: Opportunities and ethical balance

In cardiology, the implications of AI adoption are profound. Algorithms now assist in ECG pattern recognition, echocardiography interpretation, and risk prediction [23,24]. Yet, the transition from research innovation to bedside application depends on clinician trust, ethical confidence, and perceived utility. Our findings show that 225 (55.3%) participants held neutral attitudes toward AI’s potential to enhance medicine. This neutrality underscores the importance of clarity and reassurance, particularly around accountability, data transparency, and clinical decision autonomy [25,26]. Embedding ethical reflection and governance principles into AI education—alongside technical skills—can help clinicians balance enthusiasm with caution.

### Institutional and system-level considerations

AI readiness extends beyond individual clinicians. Institutional investment in training infrastructure, governance frameworks, and technical support is essential for sustainable adoption. The majority of respondents favored formal institutional AI training, echoing findings from global reviews emphasizing that organizational endorsement is a decisive factor in successful AI implementation [27,28]. Institutions should integrate AI training within continuing professional development programs, supported by policies ensuring equitable access to resources and accountability mechanisms.

### Limitations

Several limitations must be considered when interpreting these findings. The cross-sectional design prevents definitive causal inferences about the relationships between educational variables and AI acceptance. The homogeneous sample composition, while reflective of current medical training demographics, limits generalizability to more diverse healthcare professional populations and different career stages. The convenience sampling approach, including recruitment through social media platforms, may have introduced selection bias toward technology-oriented participants, potentially overestimating acceptance levels and learning motivation. The single-centre geographical focus limits generalizability to other healthcare systems, cultural contexts, or regulatory environments. Finally, reliance on self-reported measures creates potential for social desirability bias. Future research should incorporate objective knowledge assessments, longitudinal designs, and broader sampling strategies.

### Policy implications

The implications for medical education extend beyond immediate curriculum modifications to longer-term considerations about professional development, continuing education, and career-long learning in an era of rapid technological change. Future research should address the limitations identified through longitudinal designs, intervention studies testing specific educational approaches, and more diverse sampling strategies that include broader demographic representation. International comparative studies would help identify cultural and system-level factors that influence AI acceptance, while qualitative research could provide deeper insights into the underlying reasons for acceptance or resistance patterns.

The integration of AI into medical education represents not merely a technological challenge but a fundamental transformation in how medical knowledge is generated, validated, and applied in clinical practice. Understanding healthcare worker attitudes and educational preferences provides a crucial foundation for navigating this transformation successfully while maintaining the essential human elements of care.

## Materials and methods

### Study design and setting

This cross-sectional survey examined healthcare workers’ attitudes, knowledge, and educational preferences regarding the utilisation of AI in cardiac diagnosis. The study was conducted across major medical centres in Riyadh, Saudi Arabia, between January and March 2025, following full ethical approval from the Institutional Review Board of Al-Imam Muhammad Ibn Saud Islamic University.

### Ethics statement

This study received full approval from the Institutional Review Board of Al-Imam Muhammad Ibn Saud Islamic University (Riyadh, Saudi Arabia). All participants provided informed electronic consent prior to participation, with explicit acknowledgment of voluntary participation and the right to withdraw at any time without consequence.

### Participants and sampling

The target population included licensed healthcare professionals directly involved in cardiac patient care within the Riyadh healthcare system. Inclusion criteria encompassed physicians at various career stages, specialized nursing staff, and other healthcare professionals with direct cardiac patient responsibilities. Exclusion criteria comprised general practitioners without cardiac specialization, medical students, and healthcare trainees below the resident level.

We employed convenience and snowball sampling through professional networks, institutional contacts, and social media platforms (particularly WhatsApp groups) commonly used by healthcare professionals in the region. Sample size calculations indicated 385 participants were needed to achieve 95% confidence intervals with a 5% margin of error. The final sample (N = 408) exceeded this requirement.

### Survey instrument

Using Google Forms, an anonymous online questionnaire comprising four sections was administered: (1) demographic and professional characteristics; (2) previous AI experience and usage patterns; (3) attitudes and perceptions regarding AI applications in cardiac care using 5- point Likert scales; and (4) educational preferences and training needs. The AI acceptance measures were adapted from established technology acceptance frameworks [14,15] and modified for medical AI contexts.

Face and content validity were established through expert panel review in medical education, cardiology, and health technology assessment. Pilot testing with 30 healthcare professionals confirmed instrument clarity and reliability. Internal consistency of the AI acceptance scale was excellent (Cronbach’s α = 0.892).

### Data collection

Data were collected via a secure electronic survey platform distributed through institutional email networks and professional social media platforms, ensuring data security and participant confidentiality while maximising reach across different work settings.

### Statistical analysis

All analyses were conducted using SPSS version 28.0 (IBM Corp., Armonk, NY, USA). The analytical approach included: (1) descriptive statistics (frequencies, percentages, means, standard deviations); (2) reliability assessment via Cronbach’s alpha, item-total correlations, and factor analysis; (3) Spearman’s rank correlations for ordinal Likert data; (4) chi-square tests with Cramér’s V for effect size assessment (small = 0.1, medium = 0.3, large = 0.5); and (5) multiple logistic regression to identify key predictors of AI acceptance using systematic variable selection with univariate screening followed by multivariable modelling.

Model performance was assessed via the Hosmer–Lemeshow goodness-of-fit test, area under the receiver operating characteristic curve (AUC), and classification accuracy. The extremely low missing data rate (0.2%) permitted listwise deletion for primary analyses. All statistical tests employed two-tailed significance criteria (α = 0.05), with interpretation emphasising effect sizes and confidence intervals.

## Conclusions

Healthcare workers in this study demonstrated moderate acceptance of AI in cardiac diagnosis, with educational factors—particularly willingness to learn and interest in institutional training—emerging as the strongest predictors of adoption, surpassing demographic or experiential variables. These findings highlight the importance of comprehensive, hands-on educational programs that build confidence, provide practical experience, and are supported institutionally. Integrating AI literacy throughout medical curricula, rather than as an optional component, is essential for preparing clinicians for AI-enhanced practice while preserving the human elements central to care. Medical education programs have both the opportunity and responsibility to design evidence-based curricula that foster motivation, competence, and trust in AI, ensuring safe and effective integration into cardiovascular care.

## Data Availability

The de-identified dataset supporting the findings of this study is available from the corresponding author (Ghaiath Hussein) upon reasonable request. Access will be granted for non-commercial research purposes, subject to institutional data sharing policies, approval from the Institutional Review Board of Imam Mohammad Ibn Saud Islamic University, and execution of a data use agreement to ensure participant privacy and confidentiality are protected. The full dataset cannot be made publicly available due to ethical restrictions aimed at protecting participant anonymity and in accordance with the study’s informed consent and institutional guidelines.

## Acknowledgments

The authors express appreciation to all healthcare workers who participated in this survey. Special thanks are extended to the medical education and technology assessment experts who provided guidance during instrument development and validation.

## Declarations

### Data availability

The datasets generated and analysed during this study are available from the corresponding author upon reasonable request, subject to institutional data sharing policies and participant privacy protection requirements.

### Author contributions

Mohammad AlShammri: Conceptualization, Supervision, Writing – original draft, Writing – review & editing. Renad Alshehri, Ghaida Almasari, Abdullah Alrashed, Raghad Alarfaj, Renad Alabdulrahman, Mohannad Aldosari: Data curation, Investigation, Methodology, Writing – review & editing. Mosfer A Al-Walah, Meteb M Alotiabi: Formal analysis, Writing – review & editing. Ghaiath Hussein: Conceptualization, Methodology, Writing – original draft, Writing – review & editing. All authors reviewed and approved the final manuscript.

### Ethics approval and consent to participate

This study received full approval from the Institutional Review Board of Al-Imam Muhammad Ibn Saud Islamic University (Riyadh, Saudi Arabia). The study was conducted according to the principles expressed in the Declaration of Helsinki. All participants provided informed electronic consent prior to participation, with explicit acknowledgment of voluntary participation and the right to withdraw at any time without consequence.

### Consent for publication

Not applicable to this study design.

